# Identifying SARS-CoV-2 Variants of Concern through saliva-based RT-qPCR by targeting recurrent mutation sites

**DOI:** 10.1101/2022.03.02.22271785

**Authors:** Rachel E. Ham, Austin R. Smothers, Rui Che, Keegan J. Sell, Congyue Annie Peng, Delphine Dean

## Abstract

SARS-CoV-2 variants of concern (VOCs) continue to pose a public health threat which necessitates a real-time monitoring strategy to compliment whole genome sequencing. Thus, we investigated the efficacy of competitive probe RT-qPCR assays for six mutation sites identified in SARS-CoV-2 VOCs and, after validating the assays with synthetic RNA, performed these assays on positive saliva samples. When compared with whole genome sequence results, the SΔ69-70 and ORF1aΔ3675-3677 assays demonstrated 93.60% and 68.00% accuracy, respectively. The SNP assays (K417T, E484K, E484Q, L452R) demonstrated 99.20%, 96.40%, 99.60%, and 96.80% accuracies, respectively. Lastly, we screened 345 positive saliva samples from December 7-22, 2021 using Omicron-specific mutation assays and were able to quickly identify rapid spread of Omicron in Upstate South Carolina. Our workflow demonstrates a novel approach for low-cost, real-time population screening of VOCs.

**Importance:** SARS-CoV-2 variants of concern and their many sublineages can be characterized by mutations present within their genetic sequences. These mutations can provide selective advantages such as increased transmissibility and antibody evasion, which influences public health recommendations such as mask mandates, quarantine requirements, and treatment regimens. Our real-time RT-qPCR workflow allows for strain identification of SARS-CoV-2 positive saliva samples by targeting common mutation sites shared between VOCs and detecting single nucleotides present at the targeted location. This differential diagnostic system can quickly and effectively identify a wide array of SARS-CoV-2 strains, which can provide more informed public health surveillance strategies in the future.

## Introduction

SARS-CoV-2 has caused more than 407 million infections and more than 5.7 million deaths globally^1^. Under neutral genetic drift conditions, SARS-CoV-2 mutates at an estimated rate of 1×10^−3^ substitution per base per year^2^. While most mutations are insignificant, some mutations provide selective advantages, such as increased transmissibility and antibody evasion^3-5^. Several emerging strains share common nucleotide substitutions at sites that may confer advantageous phenotypic traits^6^ and have been deemed variants of concern (VOC) by public health authorities^7^.

The gold standard for differentiating variants of SARS-CoV-2 is whole genome sequencing, which provides excellent resolution of genetic information. However, for timely clinical diagnostic applications, such as real time population surveillance and treatment recommendations, using whole genome sequencing is less feasible because it is not routinely performed in clinical laboratories^9^. Additionally, diagnostic sequencing is limited by slow turnaround times and high cost per sample^10^. This necessitates a low-cost strategy for population-level surveillance of SARS-CoV-2 variants.

RT-qPCR has been used to detect population-level spread of SARS-CoV-2 VOCs, including Alpha (B.1.1.7), Beta (B.1.351), Gamma (P.1), and Delta (B.1.617.2). Alpha was initially traced through populations via S gene target failure^11^. This prompted researchers to design assays that rely on target gene failure for detection of deletions or single nucleotide polymorphisms (SNPs) in VOCs^12,13^. However, RT-qPCR assays featuring competitive probes for both reference and mutation sequences increases specificity, providing a more robust strain typing panel. Such assays have been used to detect Spike (S) deletion 69-70 along with several SNPs characteristic of Alpha and Gamma^14^. Additionally, commercially available Spike SNP assays have been used to detect Alpha, Beta, Gamma, and Delta from specimens originating from hospitalized individuals^15^. While these assays have been validated for extracted RNA originating from nasopharyngeal swabs, little work has demonstrated the efficacy of RT-qPCR VOC detection in saliva. Saliva-based RT-qPCR has been established as an accurate diagnostic tool comparable to traditional nasopharyngeal swab tests^16-20^ and warrants examination as a SARS-CoV-2 VOC detection strategy.

Many VOCs contain advantageous genotypes that have emerged independently, indicating that mutation site assays are an effective strategy to monitor emerging dangerous strains^21^. We chose to evaluate assays for biochemically significant mutations that also provide differential strain typing for SARS-CoV-2 VOCs, namely SΔ69-70, ORF1aΔ3675-3677, K417T, E484K, E484Q, and L452R. We designed an in-house assay for SΔ69-70, which has been associated with enhancement of other Spike receptor binding domain (RBD) mutations to increase infectivity in strains such as B.1.1.7^22^. We also designed an assay for ORF1aΔ3675-3677; although it has not been experimentally linked to improved viral fitness, it has been used to differentiate between Beta and Gamma VOCs^12^. We also evaluated the efficacy of TaqPath assays for K417T, E484K, E484Q, and L452R in saliva. Computational modeling has indicated that RBD residues K417, E484, and L452 are critical for increasing viral binding affinity to host cell receptors^23^. K417T and K417N SNPs^24^ and many substitutions at E484^25^ also reduce viral susceptibility to neutralizing antibodies. Additionally, L452R increases both structural stability and viral fusogenicity, and decreases cell-mediated immune response^26^. Conveniently, the currently circulating Omicron variant (B.1.1.529) harbors both L452R and SΔ69-70, so we used these assays to quickly identify its emergence at Clemson University and the surrounding Upstate South Carolina in December 2021. All assays were validated via comparison against whole genome sequence results.

## Results

### Analytical Sensitivity and Efficiency of Mutation Site-Specific RT-qPCR Assays

We evaluated the sensitivity of the mutation site-specific RT-qPCR assays via serial 10-fold dilutions of SARS-CoV-2 synthetic RNA of characteristic strains (B.1, B.1.1.7, B.1.351, P.1, B.1.617.1, B.1.617.2). The dilution range for all assays was 4⍰10^0^ to 4⍰10^6^ genome copies/assay (Table 1). We calculated RT-qPCR efficiency for both mutation and reference probes using the equation: E = -1+10^(−1/slope)^. Efficiencies of the mutation probes ranged from 89.52% to 112.04% (Table 1, other data included in Supplemental Table 1). R^2^ values for all mutation probes were ≥0.9927. The limit of detection (LoD) for SΔ69-70 was 40 genome copies/assay. LoDs for ORF1aΔ3675-3677, K417T, E484K, E484Q, and L452R were 4 genome copies/assay. LoD for the control gene (N gene) was also 4 genome copies/assay (Supplemental File 1), which is comparable to the range of detection for saliva-based clinical assays for SARS-CoV-2 screening^28^.

**Table 1.**
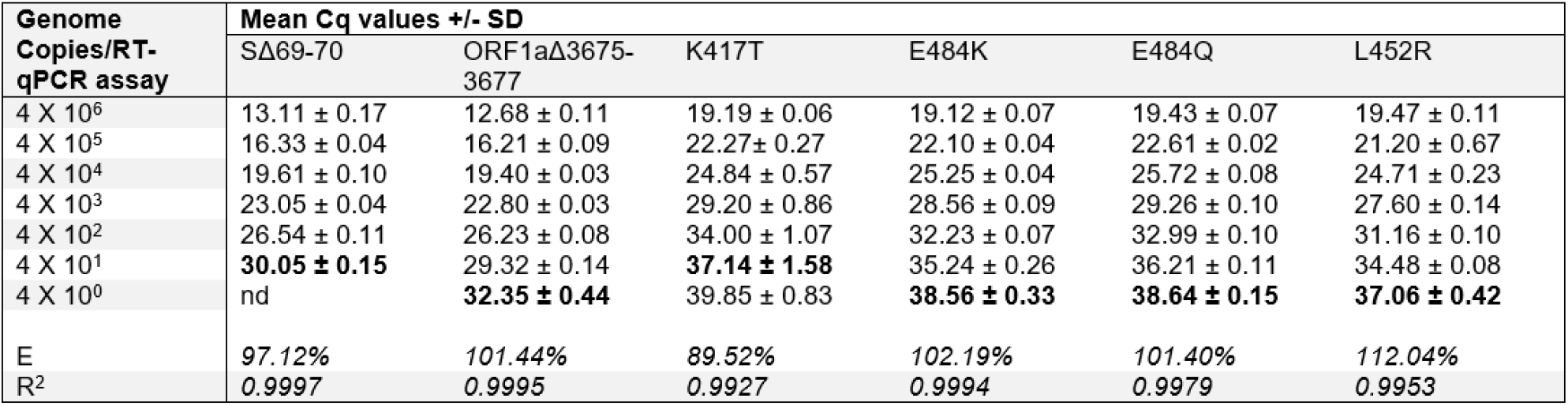
Performance of RT-qPCR deletion assays in saliva. Limits of detection are in bold.

### Analytical Specificity of Deletion Assays and Comparison with Saliva Samples

We assessed analytical specificity by performing SΔ69-70 and ORF1aΔ3675-3677 deletion assays on synthetic RNA from six characteristic SARS-CoV-2 strains at 4⍰10^4^ genome copies/assay (Figure 1). We did not observe cross-reactivity or amplification failure for any synthetic RNA on either assay. However, the deletion probe from ORF1aΔ3675-3677 produced low fluorescent output. We observed a wide range of fluorescent output from sequenced positive saliva samples (n=125) on both deletion assays. For both deletion assays, samples with low viral copy number (determined by N1 Ct values) were more likely to produce results that could not be resolved. This was especially observed in samples with N1 Ct > 25; 9.09% of samples above this threshold failed on SΔ69-70, while 27.27% of samples above this threshold failed on ORF1aΔ3675-3677 (Supplemental File 2).

**Figure 1.**
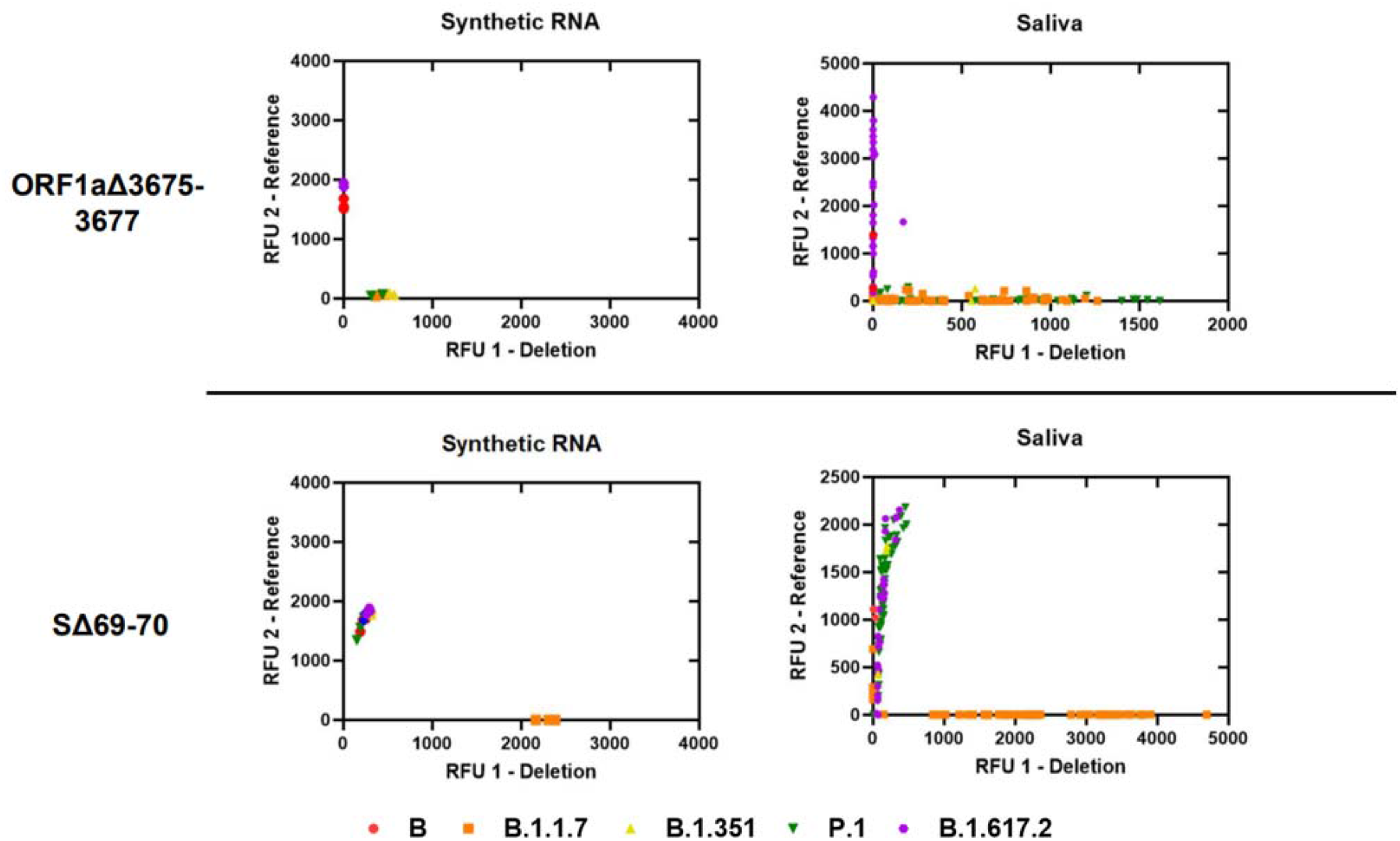
Allelic discrimination plots of deletion assays ORF1aΔ3675-3677 and SΔ69-70. Synthetic RNA controls from six SARS-CoV-2 type strains was amplified in triplicate at 4⍰10^4^ genome copies/assay via TaqPath RT-qPCR along with no template controls. The deletion probe from the ORF1aΔ3675-3677 assay produced low intensity fluorescence. Sequenced positive saliva samples (n=125) were loaded in duplicate to determine the detection range of the assay in saliva. Data were plotted by using the absolute fluorescence of each reporter dye probe.

### Analytical Specificity of Spike SNP Assays and Comparison with Saliva Samples

We assessed analytical specificity by performing K417T, E484K, E484Q, and L452R assays on synthetic RNA from six characteristic SARS-CoV-2 strains at 4⍰10^4^ genome copies/assay (Figure 2). We did not observe cross-reactivity for any synthetic RNA on any assay. Amplification failure was expected and occurred for strains lacking both reference and mutation sequences at the locus (e.g., B.1.351 lacks both alleles at K417T, B.1.351 and P.1 lack both alleles at E484Q, and B.1.617.1 lacks both alleles at E484K) which indicates high specificity of all assays performed on synthetic RNA. In saliva, we observed tight clustering of fluorescent output from sequenced positive samples (n=125) on all SNP assays. Furthermore, of the 96 replicates that produced an inconclusive result, 74 were due to the presence of an alternate allele: 70 replicates containing E484K (B.1.351 and P.1 lineages) were inconclusive on E484Q, 4 replicates containing K417N were inconclusive for K417T (B.1.351 lineage and AY.2 sublineage) (Supplemental File 3).

**Figure 2.**
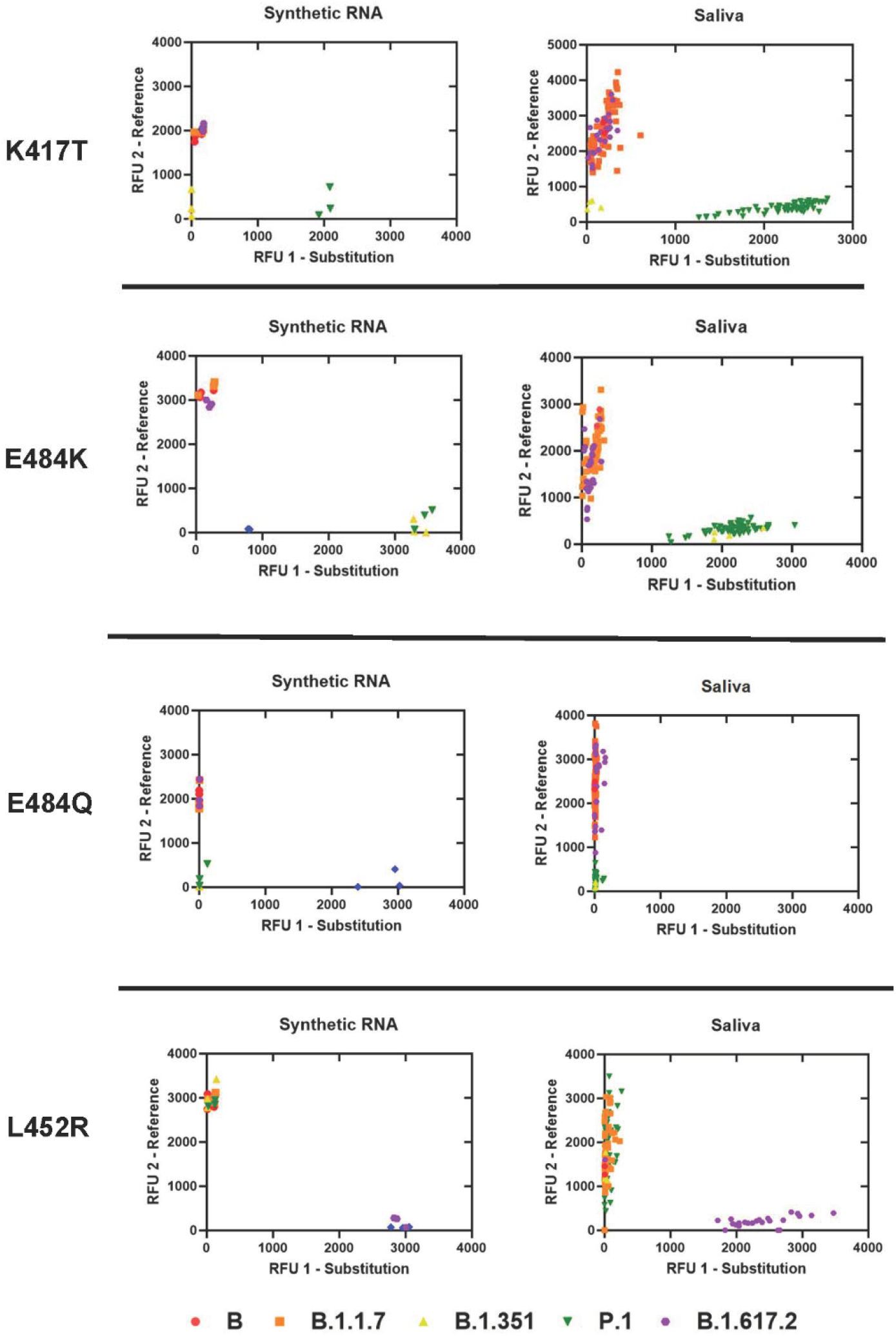
Allelic discrimination plots of SNP assays for Spike K417T, E484K, E484Q, L452R. Synthetic RNA controls from six SARS-CoV-2 type strains was amplified in triplicate at 4⍰10^4^ genome copies/assay via TaqPath RT-qPCR along with no template controls. Synthetic RNA strains that failed to amplify on K417T, E484K, E484Q, or L452R assays lacked both alleles. Sequenced positive saliva samples (n=125) were loaded in duplicate to determine the detection range of the assay in saliva. Data were plotted by using the absolute fluorescence of each reporter dye probe.

### Clinical Performance of Deletion Assays and Spike SNP Assays in Saliva

We compared assay results with whole genome sequence results to determine clinical sensitivity and specificity (Tables 2 and 3). True negatives and true positives are defined as correctly called reference and mutation sequences, respectively. False negatives are defined as incorrectly called reference sequences when the mutation sequence is present, and false positives are defined as incorrectly called mutation sequences when the reference sequence is present. Samples that produced N1 Ct values beyond the limit of detection were considered invalid. For each assay, sample results with allele-specific Ct values above the assay limit of detection were considered inconclusive. Furthermore, due to possible non-specific binding in the SNP assays, sample results with relative fluorescent output (RFU) values outside of the 99% confidence interval (95% for L452R) of allele-specific RFU were also considered inconclusive (Supplemental Files 4 and 5, logic shown in Supplemental Figure 3).

**Table 2.**
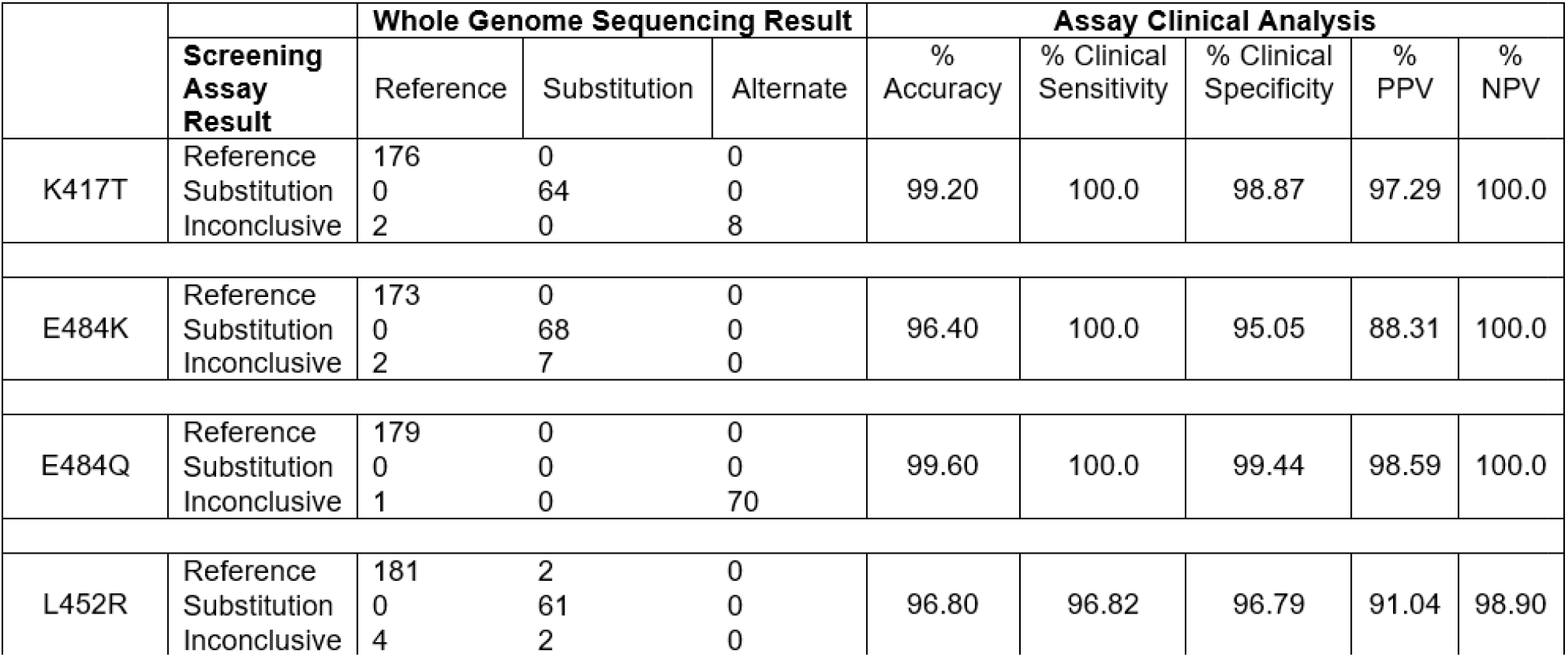
Performance of deletion assays in saliva.

**Table 3.**
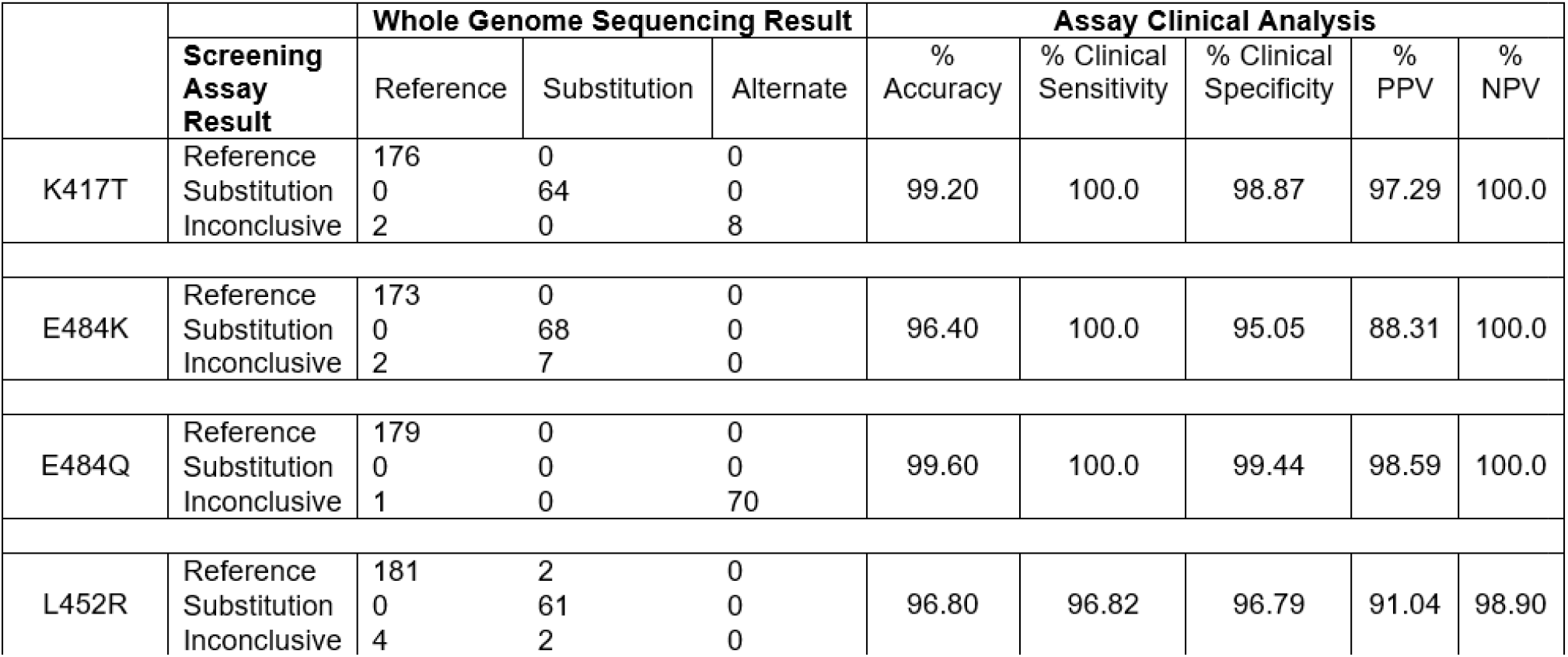
Performance of Spike SNP assays in saliva.

We calculated the total accuracies, individual probe accuracies, clinical sensitivity and specificity, as well as positive and negative predictive values for the deletion assays (Table 2). The total accuracy of SΔ69-70 was 93.6%; 92.68% of reference sequences and 96.49% of deletion sequences could be identified with the associated probes. For ORF1aΔ3675-3677, the total accuracy was 68%; 85.95% of reference sequences and 51.16% of deletion sequences could be identified with the associated probes. Clinical sensitivity was 94.82% and 95.65% for SΔ69-70 and ORF1aΔ3675-3677, respectively. Clinical specificity for both deletion assays was 100.0%. The N gene Ct values from the deletion assays indicated comparable viral loads even after the samples were stored at -80°C for over six months (Supplemental File 2). We also calculated the total accuracies, clinical sensitivity and specificity, as well as positive and negative predictive values for the Spike SNP assays in saliva (Table 3).

### Presumptive Strain Identification of Positive Saliva Samples

Combinatorial results of the six mutation sites we investigated can produce signature identification patterns for each SARS-CoV-2 VOC (Figure 3A). Consequently, we created a clinical workflow for differential strain typing based on these mutation sites (Figure 3B). Following assay validation, SARS-CoV-2 positive saliva samples were obtained from December 7-16, 2021 (n=162). Based on current circulating strains, we performed the L452R assay and identified 13 samples with reference sequence at this site. We performed the SΔ69-70 assay on these 13 samples and identified 11 with the deletion. All 13 samples were sequenced as previously described and confirmed to be B.1.1.529 (Omicron). We also screened 183 additional samples from December 17-22, 2021 to estimate prevalence of Omicron and identified 107 prospective Omicron-positive saliva samples (Supplemental File 6).

**Figure 3.**
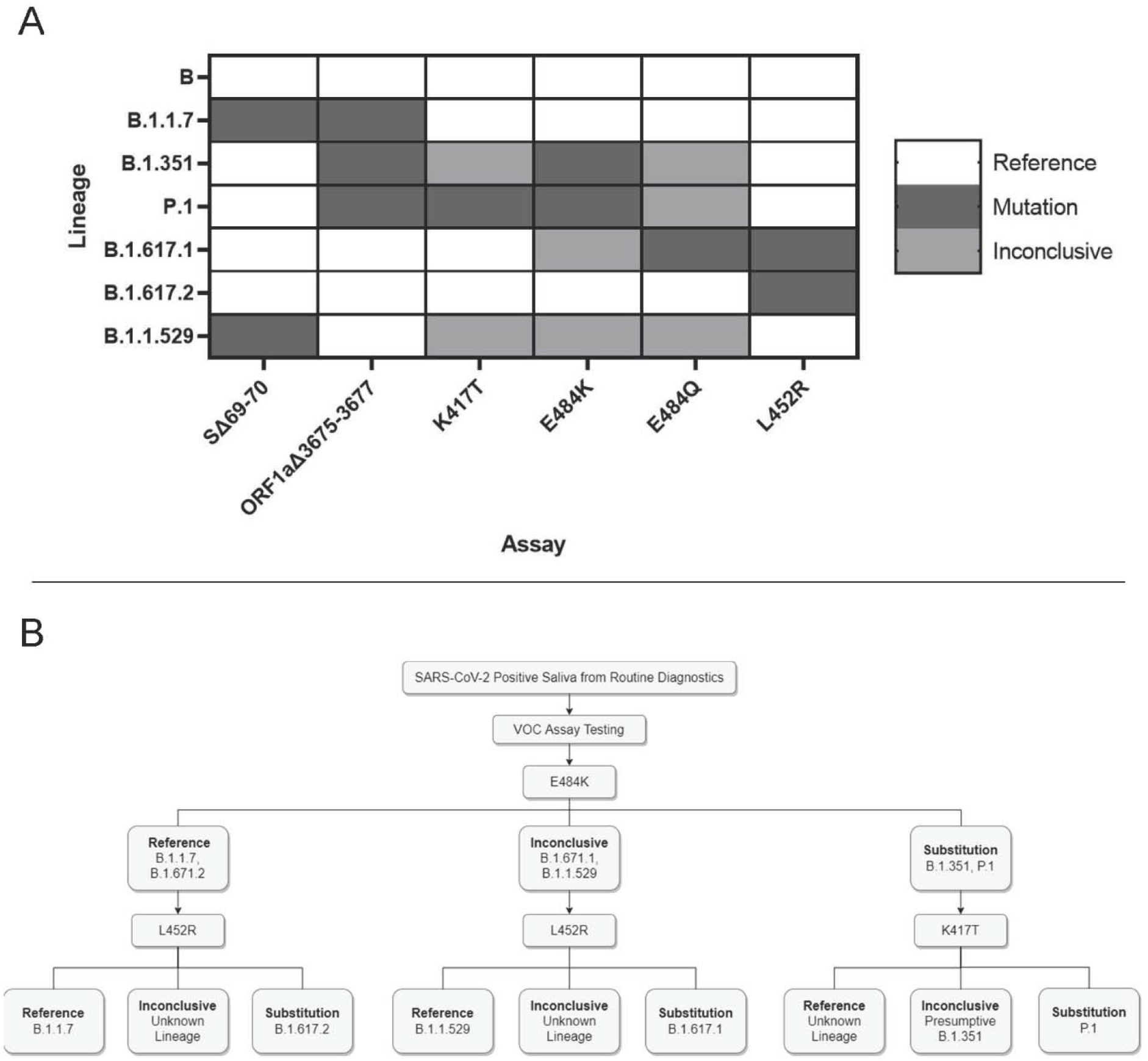
Application and interpretation of differential VOC assays. **3A**. VOC strain typing by mutation site. Each strain will produce a different combination of results from the six assays. Strains with an alternate allele at the mutation site will produce inconclusive results. **3B**. Example strain typing workflow using minimal steps. Saliva samples that are determined positive by routine diagnostic testing are analyzed by various assays that produce differential results for each VOC.

## Discussion

SARS-CoV-2 VOCs continue to pose a significant threat to public health in the United States, especially with the rapid spread of Delta starting in March 2021^29^ and, most recently, Omicron in early December 2021^30^. High transmission rates and the related clinical outcomes of these VOCs necessitate affordable and expeditious public health surveillance strategies. The lack of adequate and efficient SARS-CoV-2 variant surveillance has hindered the evaluation of clinical outcomes related to VOCs^31^. To address these limitations, our lab implemented a simple VOC screening method following our established saliva-based SARS-CoV-2 testing procedure^28^. We perform weekly surveillance testing of the entire population at Clemson University^32^, which allows for real-time monitoring of current and future VOCs.

### Deletion and SNP Assay Analyses

We designed two standard RT-qPCR assays for the SARS-CoV-2 deletion sites SΔ69-70 and ORF1aΔ3675-3677. The accuracy of the SΔ69-70 assay in saliva was 93.6% (Table 2). Two B.1.1.7 samples produced false negative results for both deletion assays, possibly due to non-specific binding of both reference probes. The accuracy of the ORF1aΔ3675-3677 assay was 68.0%; 85.9% of reference sequences could be successfully identified with the reference probe, but only 51.1% of deletion sequences could be successfully identified with the deletion probe. We believe this is due to the relative fluorescence intensity from the competitive probe pair rather than binding affinity or reaction efficiency, as both probes produced replicable amplification from synthetic RNA. The reference probe was tagged with the SUN fluor^33^, which produces high fluorescent output that can prevent the thermocycler from identifying amplification from the weaker Cy5-tagged deletion probe. We attempted to account for this by adjusting probe mixing ratios, but this did not improve the fluorescent output of the Cy5 probe.

Altering the probe pairing would likely improve the efficacy of this assay. We also performed both assays using the Luna One-Step RT-qPCR System (New England Biolabs, Ipswich MA, USA) but were unable to detect signal from the SUN probe for ORF1a3675-3677 reference sequence.

We validated four TaqPath Spike SNP assays in saliva for SARS-CoV-2 substitution sites. The accuracies of K417T, E484K, E484Q, and L452R assays were ≥ 96.4% (Table 3). Even in low viral load samples, saliva does not confound the fidelity of the assays, as the targeted sequences were accurately identified. Moreover, none of the competitive probes produced amplification at sites with an alternate allele. We did observe low-level amplification from off-target binding in the E484K, K417T, and L452R assays. High concentrations of minor grove binding probes can produce background fluorescent signal^34^. We could not further investigate if the probe concentration ratio was causing background signal because the commercial kits were premixed. However, because the assays produced well-separated signal clusters, a genotype could still be determined for samples with low-level off-target amplification.

### Probe Detection Parameters and Analysis

We determined the Ct cutoff value for each deletion and SNP assay using the absolute quantification approach estimated from analytical sensitivity^35^ and observed inconsistencies in the deletion assay limits of detection. In the SΔ69-70 assay, the internal control probe passed the Ct threshold earlier than the deletion probe across the ten-fold dilution series except for the lowest dilution, which showed a reversed relationship (4⍰10^0^ genome copies) (Supplemental File 1). Therefore, we opted to use the lower deletion probe Ct cutoff for the SΔ69-70 assay in saliva. Additionally, both assays were more likely to produce inconclusive results in saliva as the viral load decreased (indicated by N1 Ct value); this effect was more evident in the ORF1aΔ3675-3677 assay (Supplemental File 2).

Limits of detection inconsistencies were not observed with the SNP assays. However, non-specific binding of the reference probes necessitated additional RFU cutoff parameters (Supplemental Figure 3). For both deletion and SNP assays, we included both a reference and mutation control to provide a suitable constant for absolute quantification of Ct values to account for technical limitations^36^. This allowed for objective regulation of the RFU cutoff parameters to minimize investigator bias.

### Public Health Surveillance Applications

We identified the presence of the Omicron VOC in Upstate South Carolina using a combinatorial RT-qPCR strain typing strategy (Figure 3), followed by whole genome sequencing for confirmation. RT-qPCR screening for VOCs provided a strain composition estimate that allowed our public health surveillance team to adjust SARS-CoV-2 testing and health recommendations in a time-sensitive manner. This would not have been possible solely relying on whole genome sequencing because of slow turnaround time and cost. Furthermore, presumptive strain identification also influenced patient treatment recommendations from our collaborating physicians. Specifically, physicians recommended sotrovimab^37^ for COVID-19 treatment, as Delta was the predominant circulating strain at the time. Our assays indicated patient samples were positive for Omicron, which is resistant to monoclonal antibody treatment^38^. This allowed for physicians to pursue other treatment avenues.

Our strategy does not currently monitor enough unique mutation sites for comprehensive strain typing; increasing the number of targeted mutation sites maximizes the potential for strain differentiation. To address this, we are validating Spike SNP assays for K417N, N501Y, and G339D. It is important to prioritize recurring mutation sites (e.g., E484) in SARS-CoV-2 VOCs^39^ to maintain time- and cost-effectiveness. Depending on cost analysis, we would also like to implement multiplexed SNP assays. This requires custom minor groove binding probes which are more expensive than the commercially available kits we validated but can allow strain assessment in a single reaction. New predictive computational tools can identify recurring mutation sites correlated to emerging strains^40,41^, which can expedite RT-qPCR test development for real-time monitoring. Following presumptive identification, whole genome sequencing of select samples should still be performed to ensure the most accurate surveillance strategy.

### Materials and Methods

#### RT-qPCR Primer and Probe Design for Deletion Assays SΔ69-70 and ORF1aΔ3675-3677

Consensus genome sequences from Alpha (EPI_ISL_710528), Beta (EPI_ISL_678597), Gamma (EPI_ISL_792683), Delta (EPI_ISL_1544014), and a reference strain (MN908947.3) were downloaded from GenBank. Sequences were aligned using ClustalW (SnapGene v.5.4.2) to confirm that the deletions were only present in VOCs. Validated primer sets designed for the N gene^27^, SΔ69-70^12^, and ORF1aΔ3675-3677 regions^12^ matched this alignment. Each assay includes three probes tagged with different fluorophores: one targeting the N gene, one targeting the reference sequence, and one targeting the deletion sequence (Table 4). Novel reference and deletion probes were designed with short sequences to prevent primer dimer formation in a multiplex assay format. All probes were double quenched to minimize noise and maximize end point fluorescence.

**Table 4.**
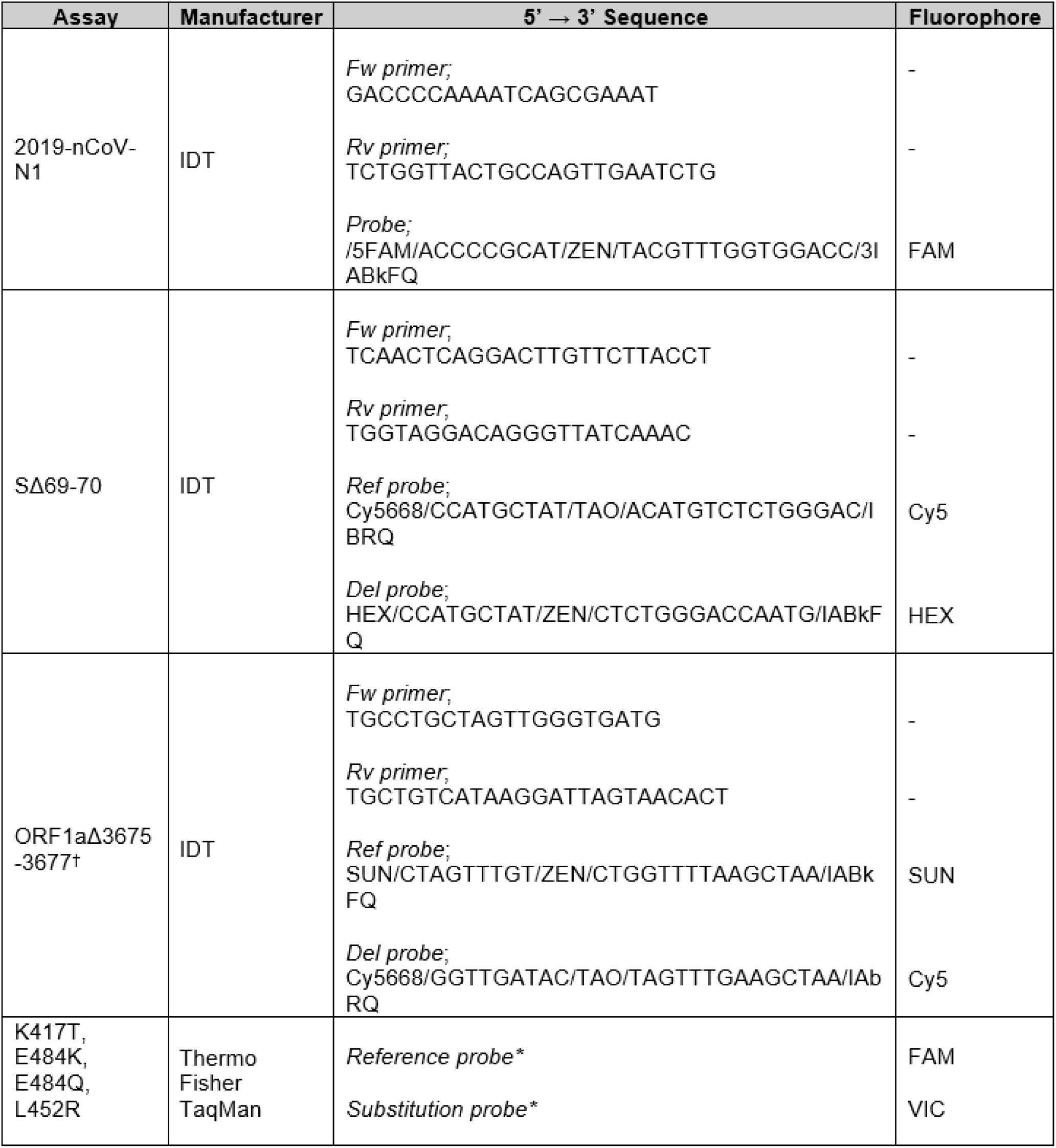
RT-qPCR probe and primer sequences for deletion and substitution assays in SARS-CoV-2 variants of concern. *sequences unavailable from the manufacturer. ^†^Open reading frame 1a.

### Optimization of Deletion and SNP Assays

SΔ69-70 and ORF1aΔ3675-3677 deletion assays were performed with TaqPath 1-Step RT-qPCR kit (Thermo Fisher, Waltham MA, USA) using reactions with 4 µL of template in a final volume of 20 µL. Primers and probes were used at final concentrations of 500 nM for each primer and 125 nM for each probe (Integrated DNA Technologies, Coralville IA, USA). SARS-CoV-2 TaqMan Assays for S substitutions K417T, E484K, E484Q, and L452R were performed per manufacturer’s instructions (Thermo Fisher) with 4 µL of template. Thermocycler conditions are described in Supplemental Table 2.

### Standard Curve and Limit of Detection Analysis

We used TWIST synthetic SARS-CoV-2 RNA control 2 (GenBank ID: MN908947.3), control 14 (GISAID ID: EPI_ISL_710528), control 16 (GISAID ID: EPI_ISL_678597), control 17 (GISAID ID: EPI_ISL_792683), control 18 (GISAID ID: EPI_ISL_1662307), and control 23 (GISAID ID: EPI_ISL_1544014) to determine the limits of detection of the screening RT-qPCR assays. We tested a 7-fold dilution series from 1,000,000 copies/μL to 1 copy/μL for both reference and mutation RNA controls in triplicate for each assay and confirmed that the lowest concentration was detected in all 3 replicates. Standard curves were created to find correlation coefficients and determine efficiencies of each probe and primer set.

### Specificity Analysis

We performed all six assays on TWIST synthetic SARS-CoV-2 RNA control 2 (B), control 14 (B.1.1.7), control 16 (B.1.351), control 17 (P.1), control 18 (B.1.617.1), and control 23 (B.1.617.2) (Twist Biosciences, San Francisco CA, USA). All synthetic RNA was diluted to 10,000 copies/μL and each reaction was performed in triplicate. Allelic discrimination plots were created for each assay to determine cross-reactivity of reference and mutation probes at each target site.

### Whole Genome Sequencing

Ethical review for this study was obtained by the Institutional Review Board of Clemson University. This study uses archived deidentified samples and data. The samples and data sets were striped of patient identifiers prior to any SARS-CoV2 sequencing and experiments for this study. Heat treated saliva samples were sequenced at a commercial lab (Premier Medical Laboratory Services, Greenville SC, USA). RNA was extracted from saliva samples via magnetic beads (Omega Bio-Tek, Norcross GA, USA) and recovered SARS-CoV-2 RNA quantity was assessed via Logix smart assay (Codiagnostics, Salt Lake City UT, USA). Samples with sufficient RNA quality were processed and sequenced on either an Illumina NovaSeq 6000 or NextSeq500/550 flow cell. Sequences were demultiplexed, assembled, and analyzed with DRAGEN COVID Lineage (Illumina, v.3.5.3).

### Saliva Screening

We performed all six assays in duplicate on sequenced saliva samples (n=125) that had greater than 95% non-N genome coverage to validate assay parameters. Saliva samples were confirmed to be SARS-CoV-2 positive and were stored at -80°C. Due to extended storage time for some positive samples, sample validity was determined using N1 Ct values to account for possible degradation. Sample identification was performed using a single-blind method and all assays were performed on all samples, removing investigator bias. We selected lineages B.1.1.7 (Alpha, n=30), B.1.351 (Beta, n=2), P.1 (Gamma, n=32), B.1.617.2/AY (Delta, n=32), and other lineages not of concern (n=29). We did not have any confirmed B.1.617.1 (Kappa, n=0) saliva samples. Five samples were excluded from analysis due to inadequate N1 amplification. GenBank accession numbers of all the sequences used to validate the assays in saliva are available in Supplemental Data.

### Statistical Analysis

We calculated accuracies ([true positives + true negatives]/sample size ⍰ 100%), clinical sensitivity (true positives/[true positives + false negatives] ⍰ 100%), clinical specificity (true negatives/[true negatives + false positives] ⍰ 100%), positive predictive value (PPV) (true positives/[true positives + false positives] ⍰ 100%), and negative predictive value (NPV) (true negatives/[true negatives + false negatives] ⍰ 100%) of the assays using whole genome sequencing results for comparison.

## Supporting information

Sequenced Sample List with GenBank and GISAID

Deletion Assay Resulting Sheet

SNP Assay Resulting Sheet

Omicron Screening Results

Standard Curve Analysis of Deletion and TaqPath Spike SNP Assays

Master Data Sheet

## Data Availability

All data produced in the present work are contained in the manuscript.

## Acknowledgements

The authors thank Clemson’s administration, medical staff, and clinical lab employees at the REDDI Lab who helped implement and manage SARS-CoV-2 testing. Thank you to Kylie King for managing the biorepository of positive saliva samples. Thank you to Dr. Mark Blenner for initial project consulting. The authors thank Jeremiah Carpenter, Kaitlyn Williams, Sujata Srikanth, and Dr. Stevin Wilson for sequencing workflow management, as well as Jessie Boulos for performing clinical sample screening. Thank you to Kylie King for technical assistance as well as critical reading of the manuscript. Thank you to Creative Inquiry students for standard curve and analytical specificity data collection. This project was funded by SC Governor and Joint Bond Review Committee, NIH NIGMS 3P20GM121342-03S1, NSF 1757658, and Clemson University Office of Creative Inquiry and Undergraduate Research.

## Author Contributions

RH, AS, and CP contributed to the conception of the manuscript. RH, AS, and RC designed project methodology. RH, RC, AS, and KS performed the experiments. RH mentored and instructed Creative Inquiry undergraduate students. RH and AS curated, analyzed, and presented data, as well as drafted the manuscript. RH, AS, DD, and CP contributed to close analysis and editing of the manuscript. All authors contributed to, read, and approved the submitted manuscript.

## Figures and Tables

**Supplemental Table 1.**
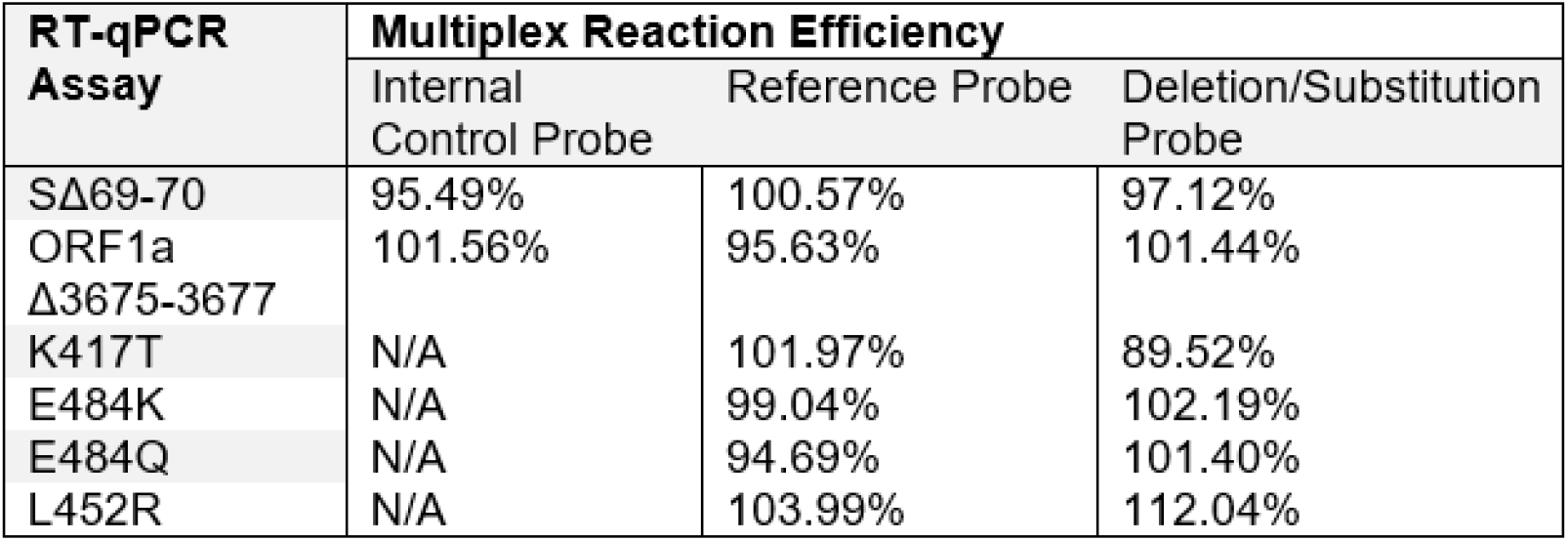
Reaction efficiencies for all primer and probe sets in multiplex.

**Supplemental Table 2.**
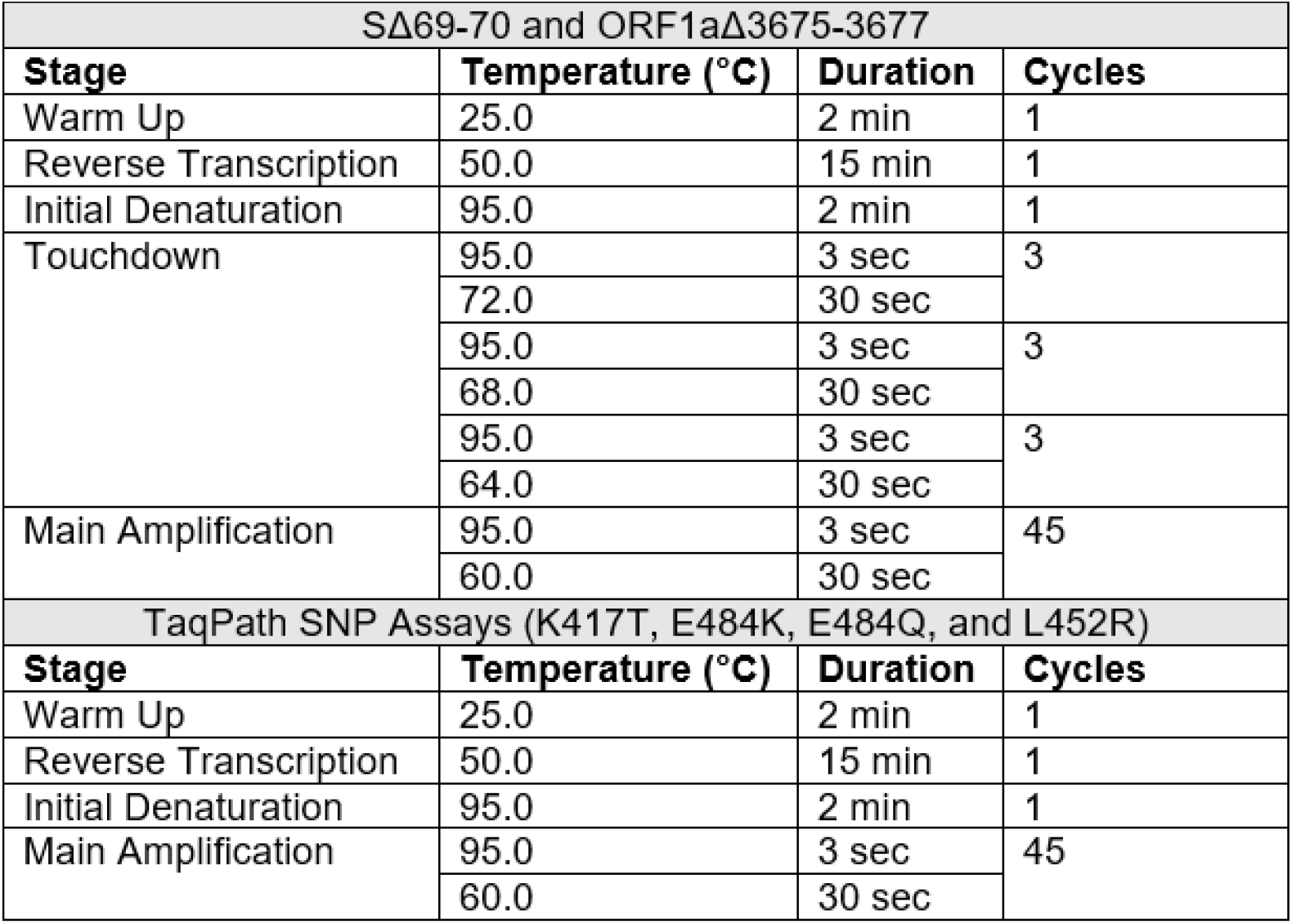
Thermocycling Conditions for RT-qPCR assays.

**Supplemental Figure 1.**
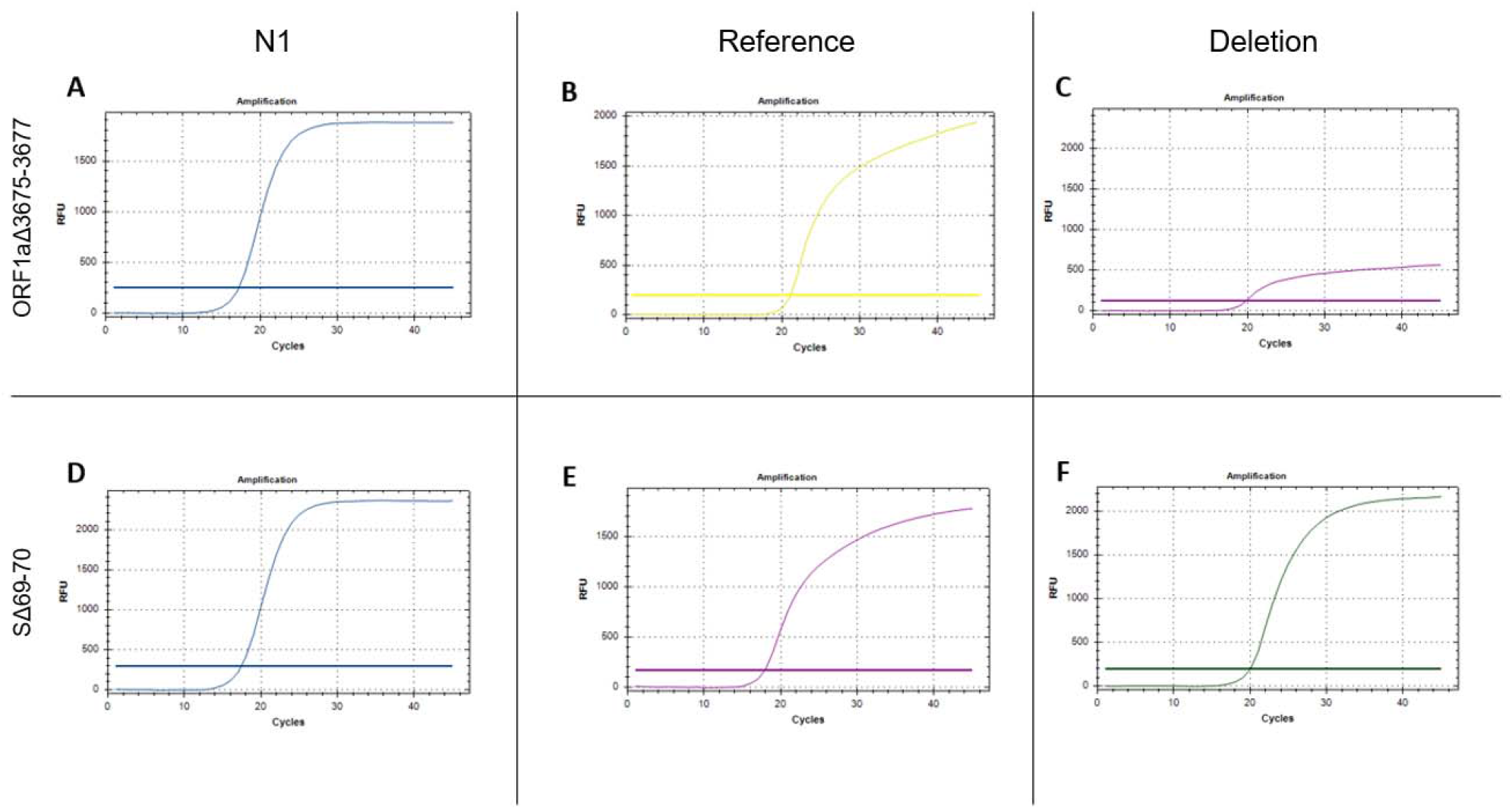
Representative curves of RT-qPCR deletion assays.

**Supplemental Figure 2.**
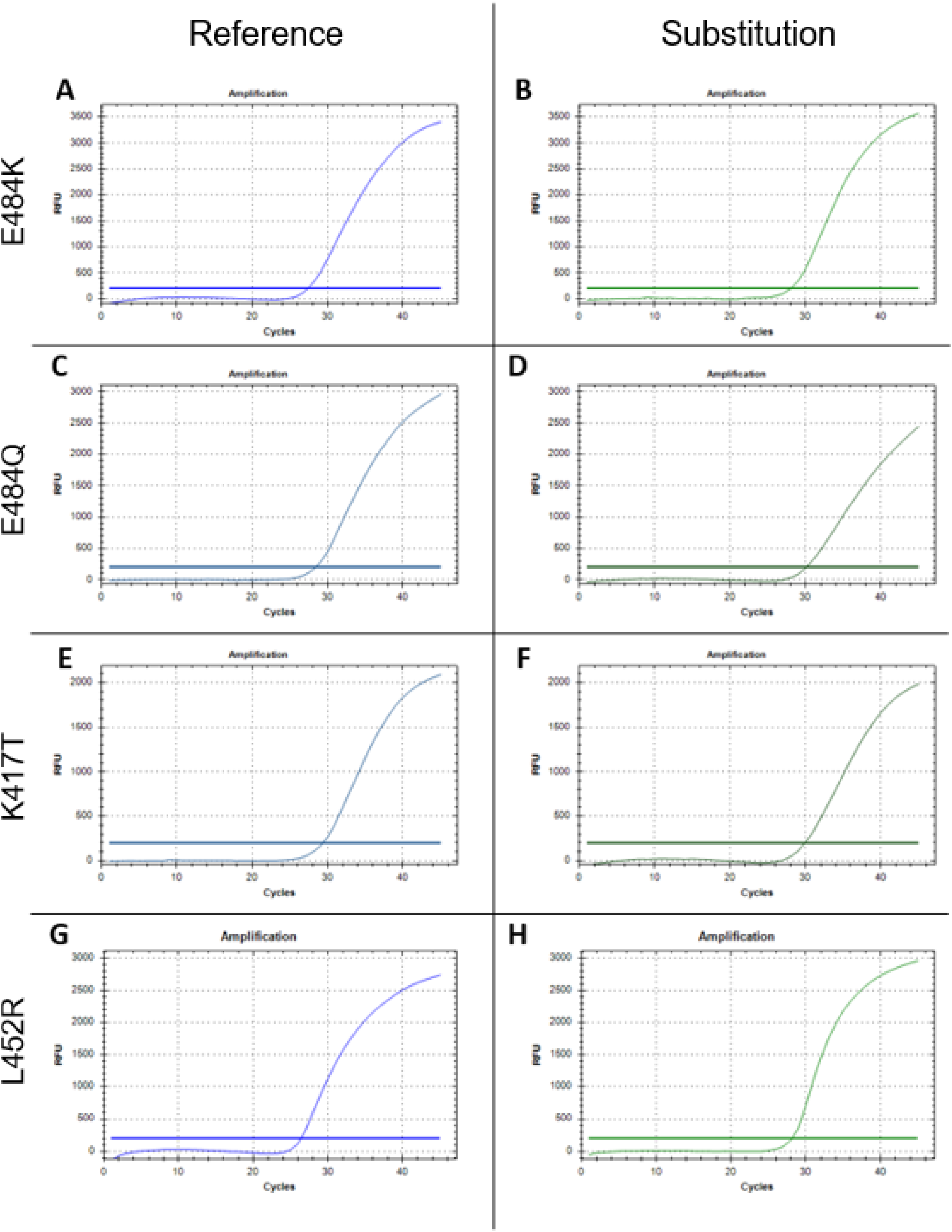
Representative curves of TaqPath Spike SNP assays.

**Supplemental Figure 3.**
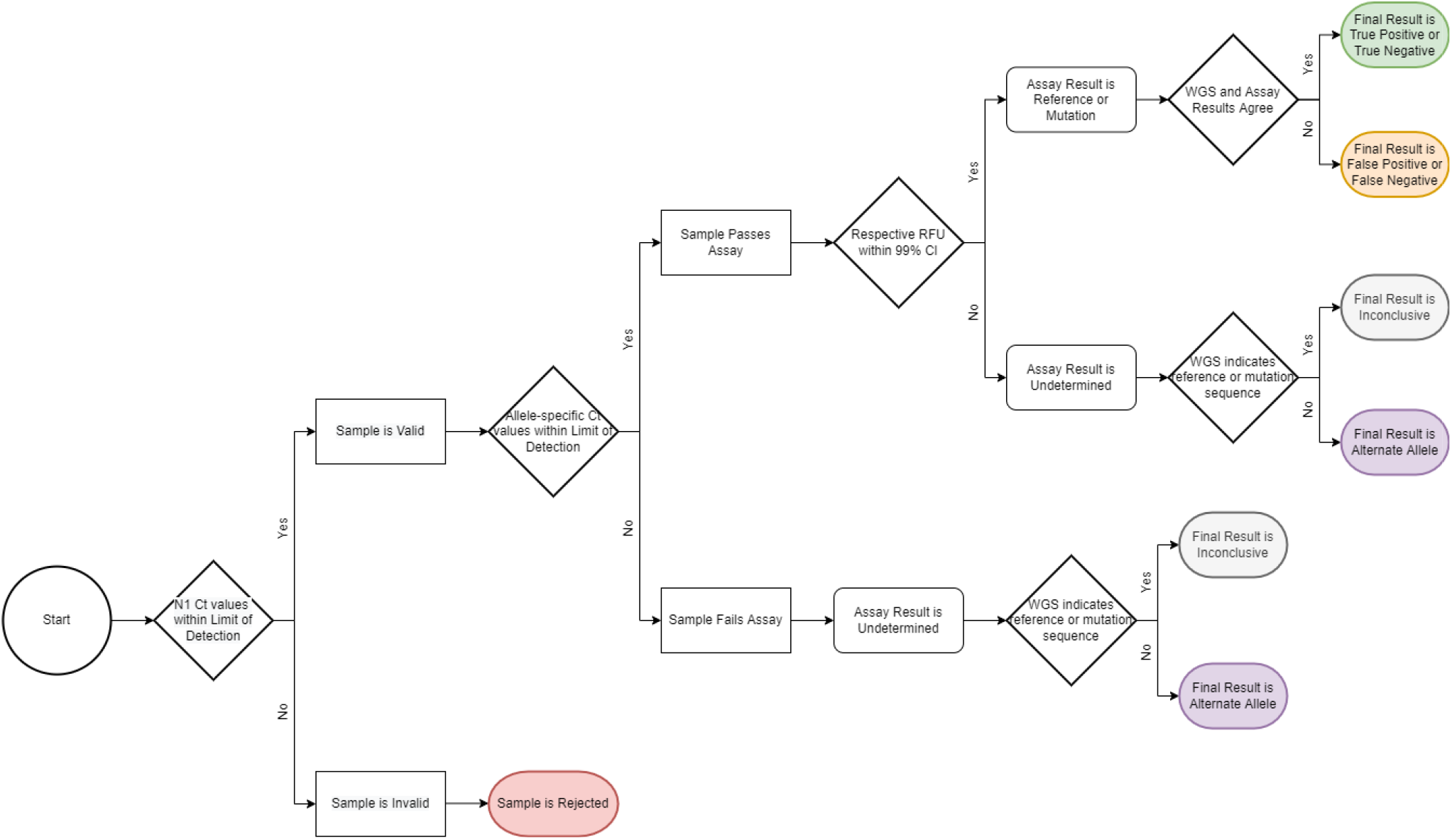
Sample Resulting Flowchart.

**Supplemental File 1**. Standard Curve Analysis of Deletion and TaqPath Spike SNP Assays

**Supplemental File 2**. Master Data Sheet

**Supplemental File 3**. Sequenced Sample List with GenBank and GISAID

**Supplemental File 4**. Deletion Assay Resulting Sheet

**Supplemental File 5**. SNP Assay Resulting Sheet

**Supplemental File 6**. Omicron Screening Results

## References

1. Dong, E., Du, H., Gardner, L. An interactive web-based dashboard to track COVID-19 in real time. Lancet Infect. Dis. 20, 533–534 (2020).

2. Duchene, S., Featherstone, L., Haritopoulou-Sinanidou, M., Rambaut, A., Lemey, P., Baele, G. Temporal signal and the phylodynamic threshold of SARS-CoV-2. Virus Evol. 6, veaa061 doi:10.1093/ve/veaa061 (2021).

3. Harvey, W.T., et. al. SARS-CoV-2 variants, spike mutations and immune escape. Nat. Rev. Microbiol., 19, 409–424 (2021).

4. Davies, N.G., et. al. Increased mortality in community-tested cases of SARS-CoV-2 lineage B.1.1.7. Nature. 593, 270–274 (2021).

5. Campbell, F., et. al. Increased transmissibility and global spread of SARS-CoV-2 variants of concern as at June 2021. Euro. Surveill., 26, 2100509 doi: 10.2807/1560-7917.ES.2021.26.24.2100509 (2021).

6. Oude-Munnink, B.B., et. al. The next phase of SARS-CoV-2 surveillance: real-time molecular epidemiology. Nat. Med. 27, 1518–1524 (2021).

7. World Health Organization. Guidance for surveillance of SARS-CoV-2 variants: interim guidance, 9 August 2021. https://apps.who.int/iris/handle/10665/343775. (2021).

8. Armstrong, G.L., et. al. Pathogen genomics in public health. N. Engl. J. Med. 381, 2569–2580 (2019).

9. Crawford, D.C., & Williams, S.M. Global variation in sequencing impedes SARS-CoV-2 surveillance. PLoS Genet. 17, e1009620 (2021).

10. Rossen, J.W., Friedrich, A.W., & Moran-Gilad, J. Practical issues in implementing whole-genome-sequencing in routine diagnostic microbiology. Clin. Microbiol. Infect. 24, 355–360 (2018).

11. Borges, V., et. al. Tracking SARS-CoV-2 lineage B.1.1.7 dissemination: insights from nationwide spike gene target failure (SGTF) and spike gene late detection (SGTL) data, Portugal, week 49 2020 to week 3 2021. Euro. Surveill. 26, 2100131 (2021).

12. Vogels, C. B. F., et. al. Multiplex qPCR discriminates variants of concern to enhance global surveillance of SARS-CoV-2. PLoS Biol. 19, e3001236 (2021).

13. Babiker, A., et. al. Single-amplicon, multiplex real-time RT-PCR with tiled probes to detect SARS-CoV-2 spike mutations associated with variants of concern. J. Clin. Microbiol. 59, e0144621 (2021).

14. Vega-Magaña, N., et. al. RT-qPCR Assays for Rapid Detection of the N501Y, 69-70del, K417N, and E484K SARS-CoV-2 Mutations: A Screening Strategy to Identify Variants with Clinical Impact. Front. Cell. Infect. Microbiol. 11, 672562 (2021).

15. Pham, V. H., et. al. (2021). Real-time PCR detects 4 rapid transmission variants of SARS-CoV-2. Int. J. Antimicrob. Agents. 58, 2100354 (2021).

16. Vogels, C. B. F., et. al. SalivaDirect: A simplified and flexible platform to enhance SARS-CoV-2 testing capacity. Med. 2, 263–280 (2021).

17. Pasomsub, E., et. al. Saliva sample as a non-invasive specimen for the diagnosis of coronavirus disease 2019: a cross-sectional study. Clin. Microbiol. Infect. 2, 285.e1-285.e4 (2021).

18. Griesemer, S. B., et. al. Evaluation of specimen types and saliva stabilization solutions for SARS-CoV-2 testing. J. Clin. Microbiol. 59, e01418–20 (2021).

19. Bastos, M. L., Perlman-Arrow, S., Menzies, D., Campbell, J. R. The sensitivity and costs of testing for SARS-CoV-2 infection with saliva versus nasopharyngeal swabs: a systematic review and meta-analysis. Ann. Intern. Med. 174, 501–510 (2021).

20. To, K. K., et. al. Consistent detection of 2019 novel coronavirus in saliva. Clin. Infect. Dis. 71, 841–843 (2020).

21. Peacock, T. P., Penrice-Randal, R., Hiscox, J. A., & Barclay, W. S. SARS-CoV-2 one year on: evidence for ongoing viral adaptation. J. Gen. Virol. 102, 001584 (2021).

22. Meng, B., et. al. Recurrent emergence of SARS-CoV-2 spike deletion H69/V70 and its role in the Alpha variant B.1.1.7. Cell Rep. 35, 109292 (2021).

23. Sanches, P. R., et. al. Recent advances in SARS-CoV-2 Spike protein and RBD mutations comparison between new variants Alpha (B. 1.1.7, United Kingdom), Beta (B.1.351, South Africa), Gamma (P.1, Brazil) and Delta (B.1.617.2, India). J Virus Erad. 7, 100054 doi:10.1016/j.jve.2021.100054. (2021).

24. Wang, L. & Cheng, G. Sequence analysis of the Emerging Sars-CoV-2 Variant Omicron in South Africa. J. Med. Virol. doi:10.1002/jmv.27516. Online ahead of print (2021).

25. Liu, Z., et. al. Identification of SARS-CoV-2 spike mutations that attenuate monoclonal and serum antibody neutralization. Cell Host Microbe. 29, 477–488 (2021).

26. Motozono, C., et. al. SARS-CoV-2 spike L452R variant evades cellular immunity and increases infectivity. Cell Host Microbe. 29, 1124–1136 (2021).

27. Centers for Disease Control. 2019-Novel Coronavirus (2019-nCoV) Real-time rRT-PCR Panel Primers and Probes. 2020 May 29. https://www.cdc.gov/coronavirus/2019-ncov/lab/rt-pcr-panel-primer-probes.html.

28. Ham, R. E., et. al. (2022). Efficient SARS-CoV-2 quantitative reverse transcriptase PCR saliva diagnostic strategy utilizing open-source pipetting robots. J. Vis. Exp. 180, e63395, doi:10.3791/63395(2022).

29. Del Rio, C., Malani, P. N., & Omer, S. B. Confronting the delta variant of SARS-CoV-2, summer 2021. JAMA. 326, 1001–1002 (2021).

30. European Centre for Disease Prevention and Control. Implications of the emergence and spread of the SARS-CoV-2 B.1.1.529 variant of concern (Omicron), for the EU/EEA. 26 November 2021.

31. Ong, S. W., Young, B. E., & Lye, D. C. Lack of detail in population-level data impedes analysis of SARS-CoV-2 variants of concern and clinical outcomes. Lancet Infect. Dis. 21, 1195–1197 (2021).

32. Rennert, L., et. al. Surveillance-based informative testing for detection and containment of SARS-CoV-2 outbreaks on a public university campus: an observational and modelling study. Lancet Child Adolesc. Health. 5, 428–436 (2021).

33. Pazdernik, N. SUN fluorophore - a molecular equivalent to VIC. 2020 June 15. https://www.idtdna.com/pages/education/decoded/article/sun-fluorophore-a-molecular-equivalent-to-vic

34. Long, S., & Berkemeier, B. Development and optimization of a simian immunodeficiency virus (SIV) droplet digital PCR (ddPCR) assay. PloS one. 15, e0240447 (2020).

35. Caraguel, C. G., Stryhn, H., Gagné, N., Dohoo, I. R., & Hammell, K. L. Selection of a cutoff value for real-time polymerase chain reaction results to fit a diagnostic purpose: analytical and epidemiologic approaches. J Vet. Diagn. Invest. 23, 2–15 (2011).

36. Wong, M. L., & Medrano, J. F. Real-time PCR for mRNA quantitation. Biotechniques. 39, 75–85 (2005).

37. Sotrovimab [package insert]. Research Triangle Park, NC: GlaxoSmithKline LLC; 2021.

38. Aggarwal, A., et. al. SARS-CoV-2 Omicron: reduction of potent humoral responses and resistance to clinical immunotherapeutics relative to viral variants of concern. Preprint at https://www.medrxiv.org/content/10.1101/2021.12.14.21267772v1 (2021).

39. Hodcroft, E. B. CoVariants: SARS-CoV-2 mutations and variants of interest. https://covariants.org/ (2021).

40. Negi, S. S., Schein, C. H., & Braun, W. Regional and temporal coordinated mutation patterns in SARS-CoV-2 spike protein revealed by a clustering and network analysis. Sci. Rep. 12, 1128 doi: 10.1038/s41598-022-04950-4 (2021).

41. Rodriguez-Rivas, J., Croce, G., Muscat, M., & Weigt, M. Epistatic models predict mutable sites in SARS-CoV-2 proteins and epitopes. Proc. Natl. Acad. Sci. 119, e2113118119 (2022).

